# Analysis of antibody data using Finite Mixture Models based on Scale Mixtures of Skew-Normal distributions

**DOI:** 10.1101/2021.03.08.21252807

**Authors:** Tiago Dias Domingues, Helena Mouriño, Nuno Sepúlveda

## Abstract

Finite mixture models have been widely used in antibody (or serological) data analysis in order to help classifying individuals into either antibody-positive or antibody-negative. The most popular models are the so-called Gaussian mixture models which assume a Normal distribution for each component of a mixture. In this work, we propose the use of finite mixture models based on a flexible class of scale mixtures of Skew-Normal distributions for serological data analysis. These distributions are sufficiently flexible to describe right and left asymmetry often observed in the distributions associated with hypothetical antibody-negative and antibody-positive individuals, respectively. We illustrate the advantage of these alternative mixture models with a data set of 406 individuals in which antibodies against six different human herpesviruses were measured in the context of Myalgic Encephalomyelitis/Chronic Fatigue Syndrome.

## 1 INTRODUCTION

Antibodies are key immunological proteins produced by B cells upon molecular recognition of an antigen derived from an infectious agent. In general, they contribute to microbial clearance and, if maintained in the body over time, they comprise the basis of the so-called immunological memory, which translates into a quicker and more efficient immune response in the case of repeated exposure to the same infectious agent. In turn, autoantibodies are antibodies recognizing components from the body and they are usually present in autoimmunity diseases, such as multiple sclerosis or rheumatoid arthritis. In the laboratory, antibodies (or autoantibodies) against a specific antigen are usually quantified by the enzymatic-linked immunosorbent assays (ELISA) using serum samples; see ref. [1, 2, 3] for some recent studies using these assays. The respective read-out is a light intensity, also known as optical density, which can be converted into a concentration or a titre using a calibration curve of known antibody concentrations. In practice, these assays are easily standardized, widely available, and ideal for high-throughput analysis of antibodies against a single antigen [1]. Such advantages make ELISA particularly suitable for large-scale sero-epidemiology surveys where one aims to estimate the prevalence of exposure to a given pathogen in the population [1, 4, 5]. With the recent development of high-throughput technologies, antibody quantification is currently shifting from the traditional ELISA to more advance assays, such as microarray [6, 7], luminex [8, 9], or cytometry bead assays [10], where a large number of different antibodies can be evaluated in the same serum sample. However, some of these promising technologies still require some degree of optimization before their widely applicability in biomedical research [11, 12].

Statistical analysis of antibody (or serological) data is usually carried out under the assumption that the antibody distribution consists of different latent populations, each one representing a distinct antibody state or different degrees of exposure to a given antigen. This assumption gives rise to a serological data analysis based on finite mixture models [13]. These models can be more or less complex depending on the number of components and mixing distributions used to describe the data. Due to its conceptual simplicity and easy of interpretation, the most popular finite mixture model in routine serological applications invokes the existence of two components related to hypothetical seronegative and seropositive individuals or, equivalently, antibody-negative and antibody-positive individuals [14, 15, 16]. Models comprising more than two components have also been found appropriate to describe data from some studies [17, 18, 19, 20, 21], but they might bring some ambiguity when interpreting which components are associated with antibody positivity [22]. In turn, the most popular choice for the mixing distributions is the Lognormal distribution in the original scale of the measurements or, equivalently, the Normal distribution after logarithmic transformation of the data [15, 17]. Gamma and Weibull are other choices for the mixing distributions among textbook probability distributions [16, 20]. Alternatively, less trivial mixture models can be used in the analysis. This is the case of a mixture between two truncated Normal distributions describing the situation where observations might fall below the lower limit of detection or above the upper limit of detection of the assay [18]. Another interesting model is a mixture between a Normal and a combination of half-Normal distributions for the hypothetical seronegative and seropositive populations, respectively [14]. The rationale behind this proposal is that the distribution of the seropositive population should be left skewed, because antibody levels tend to decrease over time [17]. Notwithstanding their suitability to tackle specific characteristics of serological data, none of the above models would appear to provide sufficiently flexibility in terms of skewness and flatnesss of each mixing distribution that could be used as the basis of data analysis automation in high-throughput serological studies.

In this scenario, we propose the scale mixture of Skew-Normal distributions (SMSN) as a flexible mixing distribution for serological data analysis. The flexibility of this family is attributed to four parameters that control the location, the scale, the skewness and the flatness of the resulting distribution. In addition, SMSN includes the Normal distribution, the Generalized Student’s t-distribution, and its skewed version as special cases [23]. We illustrate the advantage of using these models by analysing a data set related to antibodies against 6 different common herpesviruses in Myalgic Encephalomyelitis/Chronic Fatigue Syndrome (ME/CFS) [24].

## 2 DATA UNDER ANALYSIS

ME/CFS is complex disease whose patients experience a long-lasting fatigue that cannot be alleviated by rest or suffer from post-exertional malaise upon minimal physical and mental activity [25, 26]. The aetiology of the disease remains unknown, but it is often linked with common viral infections, including common herpesviruses [27].

To accelerate current knowledge on ME/CFS, it was created a large disease-specific biobank in the United Kingdom [28, 29]. The data set under analysis is part of this biobank and it was published in a recent study with the aim of investigating the immunological component of the disease [24]. In the data set, there is a total of 406 individuals, all adults, divided into three main groups: healthy controls (HC, *n* = 107; 26.4%), patients with ME/CFS (*n* = 250; 61.8%), and patients with multiple sclerosis (MS, *n* = 49; 12.1%). The group of patients with ME/CFS was further divided into a subgroup of 196 patients with mild or moderate symptoms (ME-M) and another subgroup of 54 severely affected patients who are home- or even bed-bound (ME-S). A detailed description about the recruitment of study participants, inclusion/exclusion criteria, and ethics can be found in the original reference [24].

The data set comprises six serological variables corresponding to the antibody concentration against the following common herpesviruses: human cytomegalovirus, CMV; Epstein-Barr virus, EBV; human herpesvirus-6, HHV-6; types 1 and 2 herpes simplex viruses, HSV-1 and HSV-2, respectively; and varicella-zoster virus, VZV. Note that the tested antibodies against EBV were specific to the viral-capsid antigen.

In each serum sample, the concentration of each vira-specific antibody was expressed in arbitrary units per ml (U/ml) according the corresponding optical density determined by commercial ELISA kits. According to the ELISA’s kit manufacturers, samples with antibodies concentration ≤ 8 U/ml should be classified as seronegative and those with concentration ≥ 12 U/ml should be classified as seropositive for all antibodies with the exception of HHV-6. Samples with IgG concentration between 8 and ≥ 12 U/ml should be classified as equivocal. For antibodies against HHV-6, seronegative and seropositivity should be defined as ≤10.5 U/ml or ≥ 12.5 U/ml, respectively. Samples with concentrations between these two limits were considered equivocal.

### 2.1 Ethical approval

All participants provided written informed consent for data collection (questionnaire, clinical measurement and laboratory tests), and for allowing their samples to be available to any research receiving ethical approval. Participants received an extensive information sheet and consent form in which there was an option for participation withdraw from the study at any time. Ethical approval was granted by the London School of Hygiene & Tropical Medicine (LSHTM) Ethics Committee (Ref. 6123) and the National Research Ethics Service (NRES) London-Bloomsbury Research Ethics Committee (REC ref. 11/10/1760, IRAS ID: 77765).

## 3 STATISTICAL ANALYSIS OF SEROLOGICAL DATA

### 3.1 Finite mixture models based on SMSN distributions

When analysing serological data related to the antibody responses against a specific antigen, it is usually assumed the existence of two or more latent, unobserved populations, which might represent different levels of exposure to that antigen. For simplicity, individuals that were never exposed to a given antigen are considered as seronegatives whilst individuals exposed to it are considered seropositives. In this scenario, the respective data from a specific antibody are typically described by a mixture of two or more probability distributions.

Let *G*_1_,…, *G*_*g*_ be the partition from a superpopulation *G* (sample space) and *π*_1_,…, *π*_*g*_ the probabilities of sampling an individual belonging to each latent population (with the usual restriction of 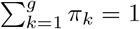 and 0 ≤ *π*_*k*_ ≤ 1). A random variable *Z* is a finite mixture of independent random variables *Z*_1_, *Z*_2_,…, *Z*_*g*_ if the probability density function (pdf) of *Z* is given by

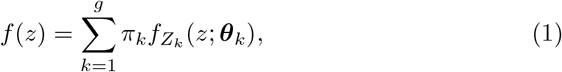

where 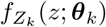 is the mixing probability density function of *Z*_*k*_ associated with the *k*-th latent population and parameterized by the vector 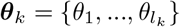.

The most popular choice for the mixing distribution in serological analysis is the Normal distribution which is symmetric around the mean and it is a mesokurtic distribution (with a kurtosis of 3 irrespective of the mean and standard deviation of the distribution). However, serological data from populations on the brink of malaria elimination show long tails and marked right asymmetry [16] in each latent population even after applying log-transformation. In such cases, one can use instead the Generalized Student t as the mixing distribution, because it has heavier tails than the Normal distribution. However, this distribution remains in the realm of the symmetric distributions. To incorporate asymmetry in the modelling, one can alternatively use the less-known Skew-Normal as the mixing distribution [30].

A random variable *W*_*k*_ has a Skew-Normal distribution with location param-eter *µ*_*k*_, scale parameter 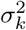 and skewness parameter *α*_*k*_ 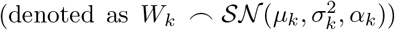 if its pdf can be written as

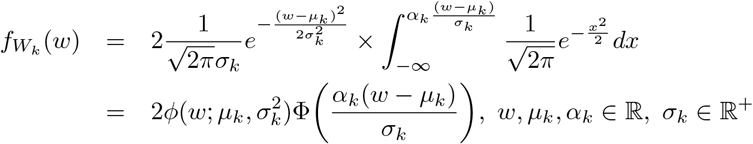

where 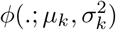 denotes the pdf of the Normal distribution with mean *µ*_*k*_ and variance 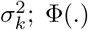 denotes the the cumulative distribution function of the standard Normal distribution [23, 31, 32]. The mean and variance of the Skew-Normal distribution are respectively given by,

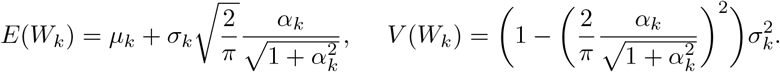

Additionally, the Skew-Normal distribution can be used to construct a more general class of flexible distributions, the scale mixtures of Skew-Normal (SMSN) distributions.

The random variable *Z*_*k*_ in expression (1) belongs to the SMSN family with location parameter *µ*_*k*_, scale parameter 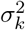 and skewness parameter *α*_*k*_ 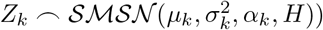 if it can be written in the following way:

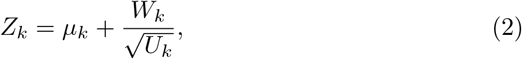

where *µ*_*k*_ is the location parameter; *U*_*k*_ is a random variable with distribution function *H*_*k*_(., ***v***_*k*_) and pdf *h*_*k*_(., ***v***_*k*_); ***v***_*k*_ is either a scalar or a vector of parameters indexing the distribution of *U*_*k*_; and 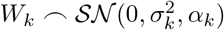 which is assumed to be independent of *U*_*k*_ [23, 33].

Based on expression (2), it is worth noting that the conditional distribution *Z*_*k*_|*U*_*k*_ = *u* takes the form

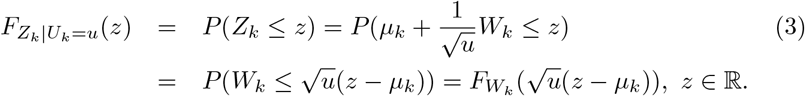

Thus,

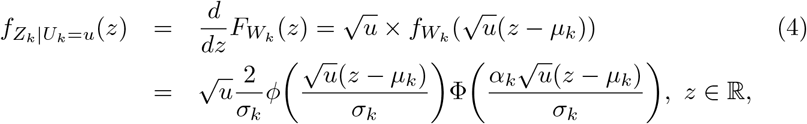

where *ϕ*(·) represents the pdf of the standard Normal distribution. Which is equivalent to, 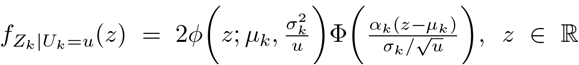, where 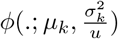 denotes the pdf of the 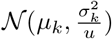. Hence, 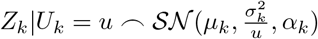.

The marginal probability density distribution of *Z*_*k*_ is given by

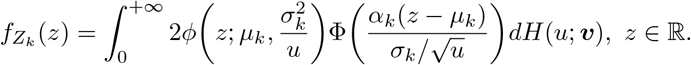

The name of this class of distributions relies on the fact that the density function of *Z*_*k*_ (2) involves an infinite mixture of Skew-Normal distributions.

To model different patterns arising from serological data, we rely on 4 particular cases of the SMSN family. The first one is the case of the Skew-Normal distribution itself. This happens when *U*_*k*_ is not a random variable but rather the scalar *u* = 1. Then, variable *Z*_*k*_ in expression (2) simplifies to *Z*_*k*_ = *µ*_*k*_ + *W*_*k*_. Hence,

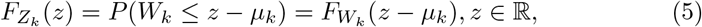

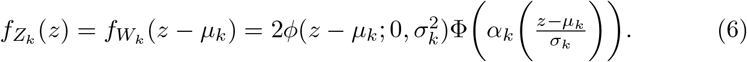

Therefore, 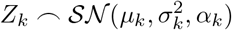.

The second case is a simplification of the previous one when *α*_*k*_ = 0. In this case, the Skew-Normal distribution reduces to the usual (symmetric) Normal distribution. In fact, when *α*_*k*_ = 0 we get

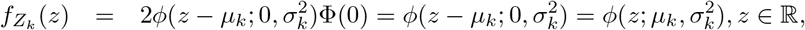

where 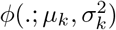 represents the pdf of the 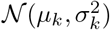 distribution.

The third and fourth cases are the skew Generalized Student’s t-distribution and its symmetric counterpart, hereafter referred to as Skew-t and Student’s t-distributions for short, respectively. These distributions can be obtained as follows.

Let *U*_*k*_ be a Gamma distribution with shape and rate parameters 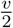 and 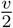,respectively, that is, 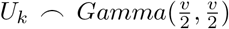. The formulation is such that the mean of *U*_*k*_ is equal to one.

Note that 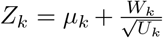, where 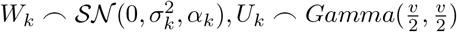 are independent random variables, is equivalent to 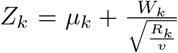 where *R*_*k*_ is a *χ*^2^ distribution with *v* degrees of freedom.

The conditional cumulative distribution function and the corresponding pdf of *Z*_*k*_|*U*_*k*_ = *u* are given by the expressions (3) and (4), respectively. According to expression (5), the marginal probability density distribution of *Z*_*k*_ takes the form

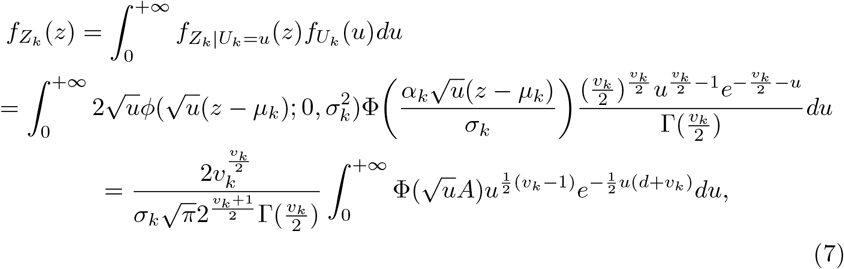

with 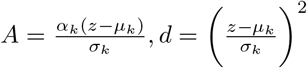.

Integrating expression (7) by substitution of the variable 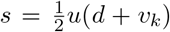, we obtain

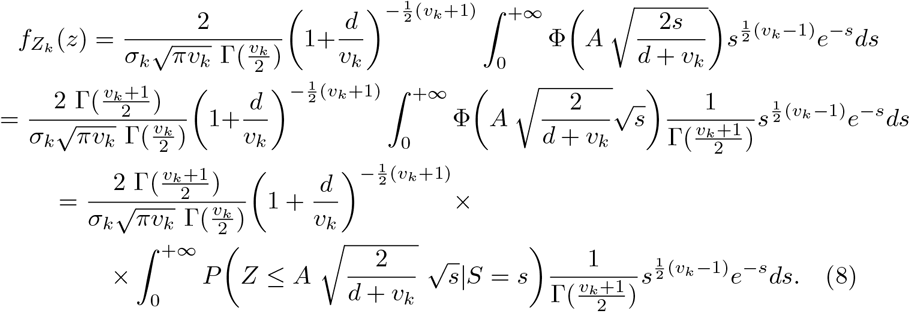

It is important to notice the following Lemma [34].

#### Lemma

Suppose that *Z*, ⌢ *𝒩* (0, 1), *Y*, ⌢ *Gamma*(*m*, 1), *R*, ⌢ *t*_2*m*_, *m >* 0. It can be proved that

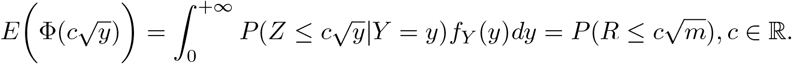

Applying this Lemma to expression (8) leads to

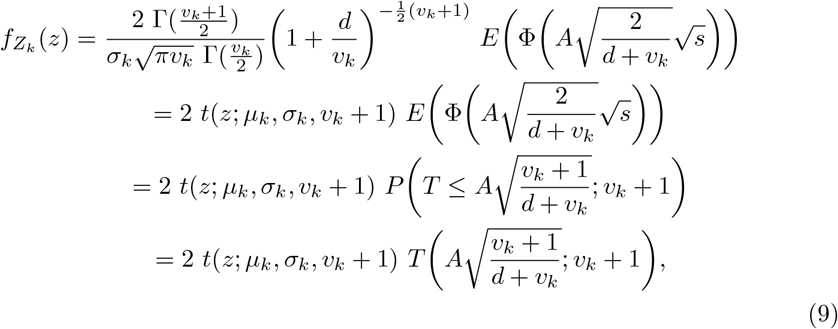

where *t*(.; *µ*_*k*_, *σ*_*k*_, *v*_*k*_ +1) denotes the probability density function of a Generalized Student-t distribution with location parameter *µ*_*k*_, scale parameter *σ*_*k*_ and *v*_*k*_ +1 degrees of freedom; *T* (.; *v*_*k*_ + 1) represents the cumulative distribution function of a standard Student-t distribution with *v*_*k*_ + 1 degrees of freedom.

In short, if 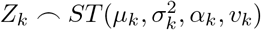, then its pdf is given by

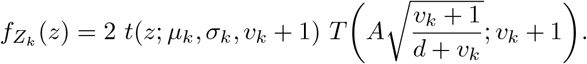

It should be noted that when the skewness parameter is equal to zero, i.e., *α*_*k*_ = 0, the quantity 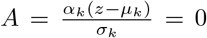, and the above expression takes the form

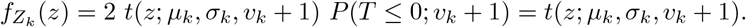

which corresponds to the probability density function of a Generalized Student-t distribution with location parameter *µ*_*k*_, scale paramter *σ*_*k*_ and *v*_*k*_ + 1 degrees of freedom.

As the degrees of freedom tends to infinity, the Skew-t distribution converges to the Skew-Normal distribution [23, 31, 32].

The mean and variance of the Skew-t distribution are respectively given by,

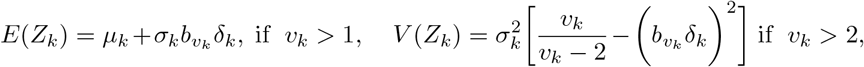

where 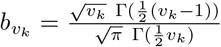 and 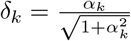.

### 3.2 Estimation and model selection

Suppose that we have a random sample *X*_1_,…, *X*_*n*_ representing the antibody levels of *n* individuals. In general, it is very difficult to determine the maximum likelihood (ML) estimates of the parameters of any given finite mixture model by direct maximization of the corresponding log-likelihood functions. One way to surpass this problem is to consider the Expectation-Maximization (EM) algorithm given that the latent serological status of each individual is unknown and, thus, we are in the presence of a problem of incomplete data.

A full derivation of an EM-type algorithm for fitting mixtures of SMSN can be found elsewhere [23]. In brief, the E-step is the same as in the traditional mixtures of Normal distributions, which has been largely studied in the literature [23, 30, 33]. Replacing the classical M-step with a sequence of conditional maximization steps (CM-steps), one obtains closed form expressions that simplify the implementation of the algorithm. Also, the observed information matrix can be derived analytically [35].

There are several methods to determine the optimal number of components that constitute the mixture, *g* [13, 36, 37]. A simple way to do it is to use penalized forms of the log-likelihood function: the information criteria. They rely on the idea that an increase in the number of components in the mixture leads to a better fit of the data, thus, increasing the maximized likelihood function. Invoking the parsimony principle to determine the best model for the data, enhancement in model fitting is penalized by an increase in the number of parameters included in the model. In this framework, two of the most popular measures are Akaike?s Information Criterion (AIC) and Bayesian Information Criterion (BIC) [38]. In general the best model is the one that provides the lowest estimate of AIC or BIC value among all models tested. About the use of one or the other criterion, the BIC criterion demonstrates consistency in determining the number of components of the mixture models [38, 39]. In addition, BIC tends to select simpler models (ideally with less number of components) than AIC, which simplifies data interpretation. Hence, this information criterion is preferred for model selection.

Another way to assess the number of components in a mixture model is to carry out a hypothesis testing, namely the Likelihood Ratio Test (LRT). However, the regularity conditions for the validity of classical asymptotic approximation of the test statistic are not met in the context of finite mixture models, because the null hypothesis associated with this hypothesis is specified in the boundary of the parameter space rather than its interior [13]. In some cases, the true parameter is in a non-identifiable subset of the parameter space [40]. As a consequence, there is no guarantee that, under the null hypothesis, the likelihood ratio statistic asymptotically follows a *χ*^2^ distribution with the degrees of freedom given by the difference between the number of parameters under the alternative and the null hypothesis [13]. To surpass this problem, a bootstrap approach can be carried out to estimate the p-value of this non-standard LRT [40, 41].

Let us consider the test specified by *H*_0_: *g* = *g*_0_ versus *H*_1_: *g* = *g*_1_ where *g*_0_ *< g*_1_. Let us also denote ***ψ***_0_ and ***ψ***_1_ the vector of parameters of the mixture models under *H*_0_ and *H*_1_ hypotheses, respectively; ***x*** = (*x*_1_,…, *x*_*n*_) the observed data and *T* (***x***; ***ψ***_0_, ***ψ***_1_) the test statistic of LRT. The bootstrap approach is given by the following algorithm [41]:

1. Use the EM algorithm to estimate the ***ψ***_0_ and ***ψ***_1_ estimates under the *H*_0_ and *H*_1_ hypotheses, respectively. Calculate 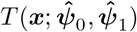;
2. Simulate *N* = 10, 000 independent samples 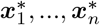 using the mixture model under *H*_0_ and parameterized by 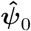;
3. For each bootstrap sample *i*, calculate 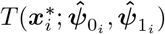, where 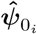 and 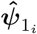 are the estimated parameter vectors for the bootstrap sample *i* under the *H*_0_ and *H*_1_ hypotheses, respectively;
4. Estimate the p-value as 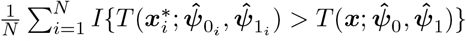, where *I* {.} is the indicator function.

Another important statistical test in the context of mixtures based on SMSN is to address the significance of the asymmetry parameters of the mixing distributions composing the mixture model based on Skew-Normal or Skew-t distributions. To attain this goal, a LRT can also be carried out. Suppose that we have a mixture model with all *g* components given by either Skew-Normal or Skew-t distributions. In this test, the hypotheses under testing are the following:

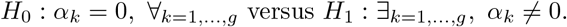

The test statistic is given by 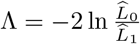, where 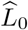 and 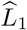 correspond to the likelihood functions of the mixture model evaluated at the maximum likelihood estimates under *H*_0_ and *H*_1_, respectively. In contrast with the previous test related to the number of components, the usual asymptotic approximation for the distribution of the LRT statistic under *H*_0_ holds, that is, a *χ*^2^ distribution with *g* degrees of freedom.

Finally, the quality of the fit of the estimated models should be assessed. For a matter of simplicity, the Pearson?s *χ*^2^ test for goodness-of-fit can be used. To apply this test, the data under analysis can be divided into bins according to the sampled 5%-quantiles or deciles (*i*.*e*., 10%-quantiles).

### 3.3 Estimation of seropositivity

After determining the best finite mixture model for the data, the next step is usually to estimate the seroprevalence, that is, the prevalence of antibody-positive individuals in the population (or, the probability of an individual being antibody-positive). Seropositivity is usually defined by a cutoff, denoted by *c*, in the respective antibody distribution above which individuals would be considered seropositive. In the context of finite mixture models, cutoff determination requires the interpretation of each latent population in terms of seronegativity and seropositivity. To do that, one typically assumes the seronegative population as the one with lowest average value while the remaining components are interpreted as different levels of seropositivity upon recurrent infections. In this scenario, the seropositivity of *i*-th individual can be seen as resulting from a Bernoulli random variable *Y*_*i*_, ⌢ *Ber*(*p*) where *p* = *P* [*X*_*i*_ ≥ *c*] and *X*_*i*_ (*i* = 1,…, *n*) represents the random variable representing the underlying antibody concentration. The probability *p* is also called seroprevalence and it embodies the probability of exposed individuals to a given antigen in the population. According to the maximum likelihood method, seroprevalence can be estimated as the proportion of seropositive individuals in the sample. Therefore, different estimates for the seroprevalence can be obtained according to the methods used to determine the cutoff.

In this work, we consider the following three different methods for determining the seropositivity cutoff:

‐ **Method 1:** It is based on the 99.9%-quantile associated with the estimated seronegative population. This method is the most popular in sero-epidemiology [22, 42]. It is often called as the 3*σ* rule, because the 99.9%-quantile is given by the mean plus 3 times the standard deviation of a normally distributed seronegative population;
‐ **Method 2:** It relies on the minimum of the density mixture functions. In the case of two latent populations, the cutoff corresponds to the absolute minimum, and in the case of three or more latent populations the cutoff corresponds to the lowest relative minimum. This point can be calculated using the Dekker?s algorithm [43]. It should be noted that the minimum of the mixing function is not expected to coincide with the point of intersection of the probability densities of each individual subpopulation;
‐ **Method 3:** It imposes a threshold in the the so-called conditional classification curves [22]. Under the assumption that all components but the first one refer to seropositive individuals, the conditional classification curve of seropositive individuals given the antibody level *x* is defined as

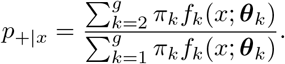

In turn, the classification curve of seronegative individuals is given by

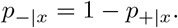

After calculating these curves, one can impose a minimum value for the classification of each individual. In this case, two cut-off values arise in the antibody distribution, one for the seronegative individuals and another for seropositive individuals. Mathematically, the classification rule is given as follows

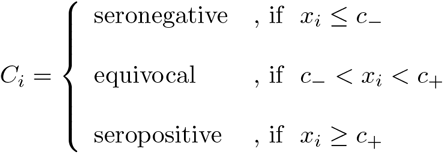

where *c*_−_ and *c*_+_ are the cutoff values in the antibody distribution that ensure a minimum classification probability, say 90%. To calculate these cutoff values in practice, one can use the bisection method providing an initial interval where they might be located [22].

### 3.4 R packages

We used the package `mixsmsn` to fit different mixture models based on SMSN [44]. In particular, we used the function `smsn.mix` to estimate the model parameter via the EM algorithm and the function `rmix` to generate random samples from a given mixture model in the bootstrap method. For fitting the Student’s t-distribution, we considered the R package `extraDistr` [45], namely, the function `dlst` to calculate their density and the function `plst` to define the cumulative distribution function. The fitting of the Skew-Normal distributions was performed with the package `sn` [46]. The functions `dsn` and `psn` were used to calculate the probability density function and the cumulative distribution function of the Skew-Normal distribution, respectively. In the case of the Skewt distribution, the functions `dst` and `pst` were used to calculate the probability density function and the cumulative distribution function, respectively.

## 4 RESULTS

### 4.1 Analysis of serological data by finite models based on SMSN

The statistical analysis was performed after applying the base 10 logarithmic transformation to the data. The number of components *g* in the mixture models was allowed to vary from 1 (single distribution) to 3 components. When fitting the mixtures of Skew-t distributions, the package `mixsmsn` only allowed to fit models with a common degree of freedom for all mixing distributions (*i*.*e*., *v*_1_ =… = *v*_*g*_ = *v*)

Our results suggested that the 6 antibodies under evaluation could be divided into two major classes: (i) the first one including antibodies against HHV-6 and VZV where there was evidence for a single serological population (Table 1) and (ii) another one including the antibodies against the remaining four herpesviruses where there was evidence for the existence of more than one serological population in the respective data (Table 2).

**Table 1:** Analysis of antibody data with evidence for a single serological population, where *g* represents the number of components in the mixture models, *p* is the number of parameters of the model, *ℒ*_max_ is the value of the maximized log-likelihood function, *p*_*gof*_ is the maximum p-value for the goodness-of-fit test when dividing data into deciles or 5%-quantiles, and *p*_*boot*_ is the bootstrap p-value for testing *H*_0_: *g* = 1 versus *H*_1_: *g* = 2.

| Virus | SMSN | $g$ | $p$ | $\mathcal{L}_{\max}$ | BIC | $p_{gof}$ | $p_{boot}$ |
| --- | --- | --- | --- | --- | --- | --- | --- |
| HHV-6 | Normal | 1 | 2 | -129.46 | 270.94 | 0.064 | 0.064 |
|  |  | 2 | 5 | -116.97 | 263.97 | 0.169 |  |
|  |  | 3 | 8 | -110.43 | 268.91 | 0.462 |  |
|  | **Skew-Normal** | **1** | **3** | **-121.35** | **260.71** | **0.140** | **0.027** |
|  |  | 2 | 7 | -117.35 | 276.75 | 0.084 |  |
|  |  | 3 | 11 | -109.40 | 284.87 | 0.152 |  |
|  | Student's t | 1 | 3 | -124.38 | 266.77 | 0.157 | 0.042 |
|  |  | 2 | 6 | -117.14 | 270.32 | 0.122 |  |
|  |  | 3 | 9 | -105.36 | 264.78 | 0.254 |  |
|  | Skew-t | 1 | 4 | -118.81 | 261.65 | 0.148 | 0.409 |
|  |  | 2 | 8 | -116.83 | 281.71 | 0.076 |  |
|  |  | 3 | 12 | -104.00 | 586.83 | 0.001 |  |
| VZV | Normal | 1 | 2 | -108.76 | 229.53 | < 0.001 | 0.000 |
|  |  | 2 | 5 | -7.28 | 44.60 | 0.159 |  |
|  |  | 3 | 8 | -1.70 | 51.45 | 0.153 |  |
|  | Skew-Normal | 1 | 3 | -23.94 | 65.90 | < 0.001 | 0.180 |
|  |  | 2 | 7 | -0.11 | 42.27 | 0.406 |  |
|  |  | 3 | 11 | 0.10 | 65.87 | 0.068 |  |
|  | Student's t | 1 | 3 | -61.90 | 141.80 | < 0.001 | 0.000 |
|  |  | 2 | 6 | -7.41 | 50.86 | 0.082 |  |
|  |  | 3 | 9 | -1.68 | 57.42 | 0.113 |  |
|  | **Skew-t** | **1** | **4** | **-7.89** | **39.81** | **0.076** | **0.375** |
|  |  | 2 | 8 | -0.05 | 48.16 | 0.211 |  |
|  |  | 3 | 12 | 5.47 | 62.14 | 0.134 |  |

**Table 2:** Analysis of antibody data with evidence for more than one serological population, where *g* represents the number of components in the mixture models, *p* is the number of parameters of the model, *ℒ*_max_ is the value of the maximized log-likelihood function, and *p*_*gof*_ is the maximum p-value for the goodness-of-fit test when dividing data in deciles or 5%-quantiles.

| **Virus** | **SMSN** | $g$ | $p$ | $\mathcal{L}_{\max}$ | **BIC** | $p_{gof}$ |
| --- | --- | --- | --- | --- | --- | --- |
| CMV | Normal | 1 | 2 | -409.11 | 830.24 | < 0.001 |
|  |  | 2 | 5 | -245.75 | 521.54 | 0.016 |
|  |  | 3 | 8 | -233.70 | 515.45 | 0.018 |
|  | Skew-Normal | 1 | 3 | -357.61 | 733.23 | < 0.001 |
|  |  | 2 | 7 | -233.82 | 509.69 | 0.038 |
|  |  | 3 | 11 | -226.64 | 519.35 | 0.146 |
|  | Student's t | 1 | 3 | -410.14 | 838.29 | < 0.001 |
|  |  | 2 | 6 | -238.54 | 513.12 | 0.038 |
|  |  | 3 | 9 | -231.23 | 516.59 | 0.046 |
|  | Skew-t | 1 | 4 | -357.71 | 739.45 | < 0.001 |
|  |  | **2** | **8** | **-231.55** | **511.45** | **0.072** |
|  |  | 3 | 12 | -226.93 | 525.93 | 0.324 |
| EBV | Normal | 1 | 2 | -342.30 | 696.62 | < 0.001 |
|  |  | 2 | 5 | -152.66 | 335.36 | < 0.001 |
|  |  | 3 | 8 | -129.30 | 306.65 | 0.173 |
|  | Skew-Normal | 1 | 3 | -226.42 | 470.86 | < 0.001 |
|  |  | 2 | 7 | -130.57 | 303.17 | 0.084 |
|  |  | 3 | 11 | -128.02 | 322.10 | 0.054 |
|  | Student's t | 1 | 3 | -240.21 | 498.43 | < 0.001 |
|  |  | 2 | 6 | -151.61 | 339.26 | < 0.001 |
|  |  | 3 | 9 | -129.41 | 312.88 | 0.117 |
|  | Skew-t | 1 | 4 | -173.14 | 370.31 | < 0.001 |
|  |  | **2** | **8** | **-125.63** | **299.32** | **0.248** |
|  |  | 3 | 12 | -126.29 | 324.66 | 0.087 |
| HSV-1 | Normal | 1 | 2 | -442.27 | 896.56 | < 0.001 |
|  |  | 2 | 5 | -291.59 | 613.22 | < 0.001 |
|  |  | 3 | 8 | -264.94 | 577.94 | 0.003 |
|  | Skew-Normal | 1 | 3 | -394.55 | 807.11 | < 0.001 |
|  |  | 2 | 7 | -260.74 | 563.52 | 0.003 |
|  |  | **3** | **11** | **-252.32** | **570.70** | **0.104** |
|  | Student's t | 1 | 3 | -443.73 | 905.48 | < 0.001 |
|  |  | 2 | 7 | -291.73 | 619.51 | < 0.001 |
|  |  | 3 | 9 | -264.98 | 584.02 | 0.002 |
|  | Skew-t | 1 | 4 | -395.43 | 814.88 | < 0.001 |
|  |  | 2 | 8 | -260.88 | 569.82 | 0.001 |
|  |  | 3 | 12 | -251.86 | 575.79 | < 0.001 |
| HSV-2 | Normal | 1 | 2 | -427.29 | 866.59 | < 0.001 |
|  |  | **2** | **5** | **-277.62** | **585.27** | **0.516** |
|  |  | 3 | 8 | -269.24 | 586.54 | 0.007 |
|  | Skew-Normal | 1 | 3 | -337.36 | 692.74 | < 0.001 |
|  |  | 2 | 7 | -264.32 | 570.68 | 0.013 |
|  |  | 3 | 11 | -257.19 | 580.45 | 0.003 |
|  | Student's t | 1 | 3 | -428.40 | 874.81 | < 0.001 |
|  |  | 2 | 6 | -277.84 | 591.71 | 0.688 |
|  |  | 3 | 9 | -269.60 | 593.26 | 0.004 |
|  | Skew-t | 1 | 4 | -337.79 | 699.60 | < 0.001 |
|  |  | 2 | 8 | -264.52 | 577.10 | 0.007 |
|  |  | 3 | 12 | -257.38 | 586.83 | 0.001 |

According to BIC, the best models for the antibodies against HHV-6 and VZV were Skew-Normal and Skew-t distributions, respectively. The estimated distributions were both left skew (Figure 1A; *α*_*HHV* 6_ = −1.82 with 95% CI =(−2.44;-−.02) and *α*_*V ZV*_ = −4.54 with 95% CI =(−6.94;−2.14). They would appear to have a good fit to the data at the 5% significance level (*p*_*gof*_ = 0.140 and 0.076, respectively). In the case of antibodies against VZV, further evidence was obtained for a single population when testing one Skew-t distribution against a mixture of two Skew-t distributions, respectively (*p*_*boot*_ = 0.375). However, when testing one Skew-Normal distribution against a mixture of two Skew-Normal distributions for the antibodies against HHV-6, the respective result was in the borderline of the 5% statistical significance (*p*_*boot*_ = 0.027).

**Figure 1.**
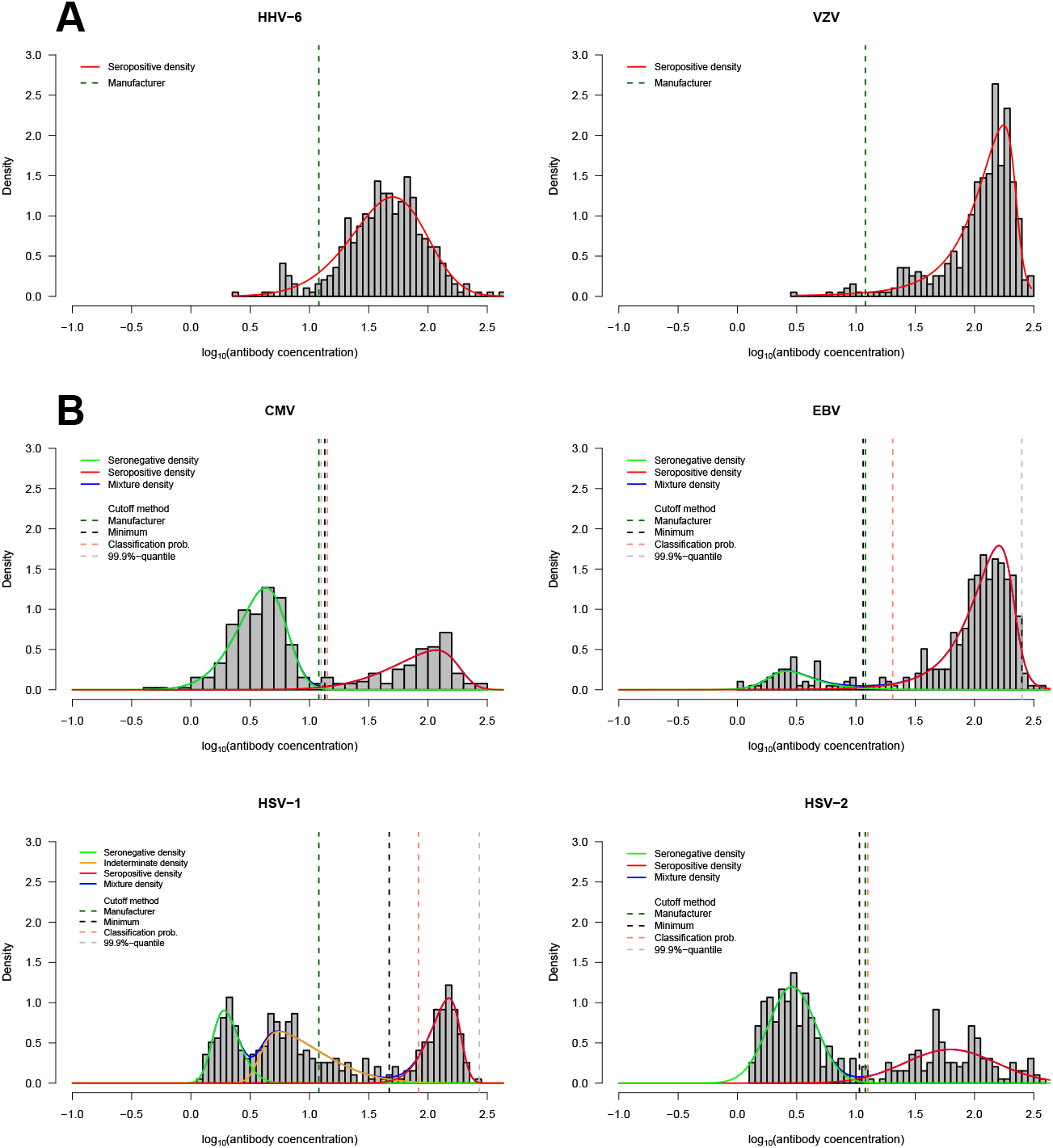
Best models for the data under analysis. **A**. Antibody distributions with evidence for a single serological population (HHV-6 and VZV). **B**. Antibody distributions with evidence for more than one serological population (CMV, EBV, HSV1, and HSV2). Antibody concentration in *x* axis is given in log_10_ units.

In terms of serological classification, the evidence for a single population would appear to represent a putative seropositive population. This interpretation is consistent with the prior knowledge that HHV-6 and VZV are usually acquired during childhood and more than 95% of the adult populations typically shows evidence of antibody positivity against these viruses [47]. In addition, the core values of these distributions are higher than the cutoff for seropositivity suggested by the lab protocol. Finally, a left skewness is also predicted for a hypothetical seropositive population, because the antibodies should decay over time [17].

It is worth noting that most of the mixture models under comparison could also fit data of these two antibodies well. This the case of the mixture of two or three Normal distributions (*p*_*gof*_ = 0.169 and 0.462 for antibodies against HHV-6 and *p*_*gof*_ = 0.159 and 0.153), which are typically used in serological data analysis. Therefore, although not being the best models for HHV-6 and VZV-related antibodies, these models could have been used for subsequent serological analyses.

With respect to the antibodies against the remaining herpesviruses, the respective data analysis was not so straightforward, because the model with lowest BIC estimate could not fit the data well according to the Pearson’s goodness-of-fit test at 5% significance level (Table 2). This is the case of the mixtures of two Skew-Normal distributions for the antibodies against CMV (BIC=509.69 and *p*_*gof*_ = 0.038), HSV-1 (BIC=563.52 and *p*_*gof*_ = 0.003), and HSV-2 (BIC=570.68 and *p*_*gof*_ = 0.013). For these antibodies, the best models were considered to be a mixture of two Skew t distributions (BIC=511.45 and *p*_*gof*_ = 0.072), a mixture of three Skew-Normal distributions (BIC=570.70 and *p*_*gof*_ = 0.104), and a mixture of two Normal distributions (BIC=585.27 and *p*_*gof*_ = 0.516), respectively, because they were the first models ranked by BIC with a good fit for the data (Figure 1B). Interestingly, for the HSV-2-related antibody data, when the mixture of two Normal distributions was compared to the mixture of 2 Skew-Normal distribution by a likelihood ratio test, the first model was strongly rejected (*p <* 0.0001), which suggested the asymmetry of at least one of the components. This inconsistency between this test and the selected model can be explained by the unavailability of fitting a mixture of a Normal distribution and a Skew-Normal distribution in the package `smsn`. For the EBV-related antibody data, the best model according to BIC was a mixture of two Skew-t distributions, which also had a good fit for the data (BIC=299.32 and *p*_*gof*_ = 0.248; Figure 1B).

In terms of interpretation of each component, there was evidence of putative seronegative and seropositive populations for antibodies against CMV, EBV, and HSV-2 (Figure 1B). This interpretation was supported by the observation that the cutoff value suggested by the commercial kits lies between these hypothetical serological populations. In the case of antibodies against HSV-1, the respective interpretation was not so obvious, because (i) the best mixture model was composed of three components and (ii) the cutoff suggested by the commercial kits lies in the middle of the intermediate distribution, which shows right asymmetry. In theory, the distribution of a putative seronegative population tends to show right asymmetry [17] and, if so, this intermediate component should be interpreted accordingly. However, this interpretation opens the door for the presence of two sets of seronegative populations resulting from distinct background signals in absence of antibodies. In absence of additional information about the serological data, this intermediate component was considered to represent a putative seronegative population.

### 4.2 Estimation of cutoff for seropositivity

After fitting the mixture models to the data, the following step of the analysis was to estimate a cutoff value for seropositivity and the subsequent seroprevalence in the different study groups (Table 3).

**Table 3:** Seroprevalence (%) by cutoff method for seropositivity and by study group. *c*_−_ and *c*_+_ are on the linear scale (U/ml). Seroprevalence was calculated based on *c*_+_. The method denoted by “M” refers to the cutoff suggested by the protocol of the commercial kit. The confidence intervals (CI) refer to the Clopper-Pearson exact confidence interval for a proportion.

| Virus | Method | $c_-$ | $c_+$ | Seroprevalence (95% CI) | | | | |
| --- | --- | --- | --- | --- | --- | --- | --- | --- |
|  |  |  |  | Global | HC | ME-M | ME-S | MS |
| CMV | M | 8.0 | 12.0 | 33.5<br>(28.9-38.4) | 37.4<br>(28.2-47.3) | 28.6<br>(22.4-35.4) | 33.3<br>(21.1-47.5) | 36.7<br>(23.4-51.7) |
|  | 1 | - | 12.6 | 33.5<br>(28.9-38.4) | 37.4<br>(28.2-47.3) | 28.6<br>(22.4-35.4) | 33.3<br>(21.1-47.5) | 36.7<br>(23.4-51.7) |
|  | 2 | - | 13.5 | 33.2<br>(28.6-38.1) | 37.4<br>(28.2-47.3) | 28.6<br>(22.4-35.4) | 31.5<br>(19.5-45.6) | 36.7<br>(23.4-51.7) |
|  | 3 | 9.4 | 14.1 | 32.9<br>(28.4-37.9) | 37.4<br>(28.2-47.3) | 28.1<br>(21.9-34.9) | 31.5<br>(19.5-45.6) | 36.7<br>(23.4-51.7) |
| EBV | M | 8.0 | 12.0 | 87.3<br>(83.6-90.4) | 87.9<br>(80.1-93.4) | 86.2<br>(80.6-90.7) | 81.5<br>(68.6-90.7) | 75.5<br>(61.1-86.7) |
|  | 1 | - | 249.5 | 2.0<br>(0.09-3.9) | 1.9<br>(0.02-6.6) | 1.5<br>(0.03-4.4) | 0.0<br>(0.0-6.6) | 6.1<br>(1.3-16.9) |
|  | 2 | - | 11.5 | 87.3<br>(83.6-90.4) | 87.9<br>(80.1-93.4) | 86.2<br>(80.6-90.7) | 81.5<br>(68.6-90.7) | 75.5<br>(61.1-86.7) |
|  | 3 | 5.6 | 20.4 | 85.5<br>(81.7-88.9) | 87.9<br>(80.1-93.4) | 82.7<br>(76.6-87.7) | 81.5<br>(68.6-90.7) | 75.5<br>(61.1-75.5) |
| HSV-1 | M | 8.0 | 12.0 | 45.2<br>(40.2-50.2) | 42.1<br>(32.6-51.9) | 41.8<br>(34.8-49.1) | 51.9<br>(37.8-65.6) | 46.9<br>(32.5-61.7) |
|  | 1 | - | 271.0 | 0.0<br>(0.0-0.1) | 0.0<br>(0.0-3.4) | 0.0<br>(0.0-1.2) | 0.0<br>(0.0-6.6) | 0.0<br>(0.0-7.3) |
|  | 2 | - | 46.9 | 34.5<br>(29.8-39.4) | 28.0<br>(19.8-37.5) | 34.7<br>(28.1-41.8) | 38.9<br>(25.9-53.1) | 34.7<br>(21.7-49.6) |
|  | 3 | 42.7 | 83.2 | 30.7<br>(26.2-35.5) | 24.3<br>(16.5-33.5) | 32.1<br>(25.7-39.2) | 33.3<br>(21.1-47.5) | 28.6<br>(16.6-43.3) |
| HSV-2 | M | 8.0 | 12.0 | 38.1<br>(33.3-43.1) | 33.6<br>(24.8-43.4) | 38.8<br>(31.9-45.9) | 40.7<br>(27.6-54.9) | 32.7<br>(19.9-47.5) |
|  | 1 | - | 12.0 | 38.1<br>(33.3-43.1) | 33.6<br>(24.8-43.4) | 38.8<br>(31.9-45.9) | 40.7<br>(27.6-54.9) | 32.7<br>(19.9-47.5) |
|  | 2 | - | 10.7 | 38.8<br>(33.9-43.8) | 33.6<br>(24.8-43.4) | 39.3<br>(32.4-46.5) | 40.7<br>(27.6-54.9) | 36.7<br>(23.4-51.7) |
|  | 3 | 7.1 | 12.6 | 37.8<br>(33.0-42.8) | 33.6<br>(24.8-43.4) | 38.8<br>(31.9-45.9) | 40.7<br>(27.6-54.9) | 30.6<br>(18.3-45.4) |

For CMV and HSV-2 antibody data, the cutoff values did not vary substantially from one method to another. Interesting, the cutoff values estimated by method 1 (the 3*σ* rule) almost perfectly matched with the ones suggested by the commercial kits (12.6 U/ml and 12.0 U/ml for CMV and HSV-2 respectively versus 12.0). This good matching between estimates could be explained by a good approximation of the Normal distribution for the seronegative population (Figure 1B) and, therefore, we could infer that the cutoff value suggested by the commercial kits was derived from the 3*σ* rule; this information was absent from the original study [24]. Since the seronegative and seropositive populations were separated well in these antibody distributions, the estimates of seroprevalence across the different study groups were almost invariant with respect to cutoff value used.

With respect to the EBV antibody data, the hypothetical seronegative population is asymmetric to the right (*α*_1_ = 1.74; 95% CI=(-−.30; 4.80); Figure 1B) with heavy tails (*v* = 4.52; 95% CI=(0.79;8.26)). As a consequence, the cut-off value of 249.5 U/ml derived from method 1 was quite different from the one suggested by the commercial kit. However, this cutoff value was considered non-informative, because it was well located within the seropositive population and implied seroprevalence estimates close to zero for the different study groups. In contrast, the cutoff values from the remaining methods were in the same order of magnitude of the one suggested by the commercial kits. Therefore, the subsequent seroprevalence estimates of each study group did not differ substantially among these methods. Again, the consistency of the resulting seroprevalence estimates was due to the fact that the seronegative and seropositive populations were well separated in these data.

The largest differences in the cutoff values for seropositivity were observed for the HSV-1 antibody data. Coincidentally, this was the data set where the best mixture model was composed of three components. As discussed earlier in this paper, the intermediate component was considered a second hypothetical seronegative population, which resulted in a shift in the calculation of seropositivity towards higher values. As such, the cutoff seropositive based on the commercial kit led to the highest seroprevalence estimates for all study groups with a global estimate of 45.2% (95% CI=(40.2%;50.2%). As an extreme case, the 3*σ* rule produced again a too-high cutoff value due to the right asymmetry of both seronegative populations. Such unrealistic cutoff value led a zero seroprevalence estimates and rendered the respective analysis useless.

Finally, although not being the main objective of this study, the comparison of the four study groups suggested that, given a method for determining seropositivity and antibody under analysis, the seroprevalence of patients with ME/CFS did not appear to differ significantly from the one of healthy controls and patients with multiple sclerosis alike.

## 5 CONCLUSIONS

This study aimed to review the finite mixture models based on SMSN and to recommend their routine use in serological data analysis. Such recommendation sets its foundation in the high flexibility of these models in describing different patterns of randomness, as illustrated with the analysis of antibodies against 6 different herpesviruses. In particular, a high modelling flexibility is necessary given that right and left asymmetry could emerge from hypothetical seronegative and seropositive populations, respectively. In this regard, most popular distributions used in statistics are not able to exhibit either left or right asymmetry depending on the parameters specified. A less-known family of distributions that shows such remarkable stochastic property is the so-called the Generalized Tukey’s *λ* distribution [50, 51]. This distribution offers a great variety of shapes owing to four parameters controlling the location, the scale, the skewness, and the flatness of the resulting distribution. However, the Generalized Tukey’s *λ* distribution is only defined in terms of its quantile function and, hence, its estimation is cumbersome. This distribution has already been proposed for mixture modelling, but there are only theoretical and computational developments available for the two-component case [48, 49]. This limits the applicability of these alternative models, namely, in data where there is evidence for more than two serological populations, such as the case of the antibodies against HSV-1 here analyzed or against the influenza virus reported elsewhere [20]. Therefore, finite mixture models based on SMSN would appear the most general and flexible approach so far for analysing serological data.

For data analysis, we recommend the use of the package `mixsmsn` for estimating the finite mixture models [44]. Notwithstanding this recommendation, the package only allows to estimate finite mixture models where all mixing distributions belong to the same class of SMSN probability distributions. Hence, it is only possible to fit 4 different models per number of components. In theory, there are 4^2^ = 16 possible two-component mixture models resulting from the combination of Normal, Skew-Normal, Generalized Student’s t, and Skew-t distributions as mixing distributions. Note that these possible models result from imposing parametric restrictions to the most general mixture model based on the Skew-t distribution. For three-component mixture models, the number of possible models increases to 4^3^ = 64. Therefore, the package `mixsmsn` excludes a vast number of possible models, which ultimately affects the detection of the true best model for the data. This computational limitation might be the reason for some inconsistencies that can be found in the example of application. For instance, a single Skew-Normal distributions was considered the best model for the antibodies against HHV-6. However, the hypothesis of a single Skew-Normal distribution against a mixture of two Skew-Normal distributions could be rejected by bootstrap at the 5% significance level. A possible explanation for this contradicting evidence is that the best model for these data could be a mixture of a Normal distribution for the seronegative population and a Skew-Normal distribution for the seropositive population.

Another limitation of using `mixsmsn` package is that, for mathematical tractability, the mixtures of generalized Student t and Skew-t distributions were assumed to have the same degrees of freedom in all the mixing distributions. In theory, this assumption could be relaxed so this parameter could vary from one component of the mixture to another. This modelling option was available in the package EMMIXuskew for the mixture of Skew-t distributions [52]. However, this package is currently discontinued. In practice, we expect some degree of numerical instability when trying to estimate different degrees of freedom for mixtures in which different components overlap with each other substantially. In this regard, future research could be conducted in order to determine under which conditions different degrees of freedom could infer for the different components. The problem of determining the optimal cutoff value for seropositivity has been intensively investigated, discussed, and revisited over the years [42, 53, 54, 55]. In this regard, the most popular cutoffs for seropositivity are simply defined by the mean plus a given number of times the standard deviation of the hypothetical seronegative population without checking the Normality assumption of the hypothetical seronegative population. The resulting cutoffs are associated with high-order quantiles of the Normal distribution, such as 97.7% or 99.9% for the 2*σ* and 3*σ* rules, respectively. In practice, these cutoffs imply a high specificity but show an arbitrary sensitivity for the respective serological classification. When the hypothetical seronegative population shows a right skew distribution, similar cutoffs can be obtained by calculating same high quantiles of the estimated SMSN, as done here. The reverse argument can be made when analysing antibodies where seropositivity is expected to be the default serological state of an individual, such as the case of antibodies against HHV-6 and VZV here analyzed or vaccine-related antibodies in populations where vaccination is mandatory. For these antibodies, similar cutoffs can be determined by the mean minus a given number of times the standard deviation of the hypothetical seropositive population assumed to be normally distributed. For a hypothetical seropositive population with a left skew distribution, the cutoff values for seropositive are now calculated using the low order quantiles (e.g., 2.3% and 0.1%-quantiles for the 2*σ* and 3*σ* rules, respectively). Inversely, these cutoffs generate a high sensitivity but an arbitrary specificity for the respective serological classification. It is worth noting that, as expected, several authors advocate a free-cutoff approach for serological analysis [15, 56]. However, a detailed discussion about the advantages and disadvantages of free-cutoff approaches was considered to be out of the scope of this study.

In terms of the results concerning the example of application, there is no evidence for a different level of exposure of the patients with ME/CFS to these herpesviruses when compared to healthy controls and patients with multiple sclerosis. This finding seems independent of the method for determining the seropositivity and it is in line with the findings reported in the original study [24] and with another serological investigation of these herpesviruses when comparing patients with healthy controls only [9]. A possible explanation for this “negative” finding might be explained by the choice of antibodies against highly immunogenic antigens used in this serological study. It is then possible that there is a specific set of viral-derived antigens associated with ME/CFS, as suggested by a comprehensive study about the role of antibodies against EBV in this disease [7]. Finally, a more detailed analysis of these data is currently carried out in order to understand whether the lack of association between ME/CFS and these antibodies could be explained by putative confounding effects of age and gender on the underlying antibody distributions. This detailed analysis will be reported elsewhere.

In summary, the finite mixture models based on SMSN show a good potential to become a routine tool for serological data analysis. They have the advantage of including the popular Gaussian mixture models as special cases. However, given the statistical complexity of these models, we recommend a closer collaboration between biomedical researchers who generate the serological data and biostatisticians who have in principle the knowledge and skills to fit and compared them properly.

## Data Availability

The data under analysis is part of a database dedicated to Chronic Fatigue, and is the responsibility of the London School of Hygiene and Tropical Medicine working group

## 6 Acknowledgments

The authors would like to thank Eliana Lacerda, Luis Nacul and Jackie-Cliff from the London School of Hygiene & Tropical Medicine (LSHTM) for sharing the data, and João Malato for helping to proof-read the manuscript. This work was partially funded by Fundação para a Ciência e a Tecnologia, Portugal (ref:UIDB/00006/2020 and UIDB/04561/2020) and resulted from a short-term scientific mission of TDS to the LSHTM funded by the EUROMENE Cost Action (CA15111) from the European Union.

## References

[1] Wang, S. S., Schiffman, M., Shields, T. S., Herrero, R., Hildesheim, A., Bratti, M. C., Sherman, M. E., Rodriguez, A. C., Castle, P. E., Morales, J., Alfaro, M., Wright, T., Chen, S., Clayman, B., Burk, R. D. and Viscidi, R. P. (2003). Seroprevalence of human papillomavirus-16, −18, −31, and −45 in a population-based cohort of 10000 women in Costa Rica, British Journal of Cancer, 89, 7, 1248-?1254.

[2] Vila Nova, B., Cunha, E., Sepúlveda, N., Oliveira, M., São Braz, B., Tavares, L., Almeida, V. and Gil, S. (2018). Evaluation of the humoral immune response induced by vaccination for canine distemper and parvovirus: a pilot study, BMC Veterinary Research, 16, 14, 348.

[3] Shikova, E., Reshkova, V. Kumanova, ?., Raleva, S., Alexandrova, D., Capo, N., Murovska, M. and European Network on ME/CFS (EUROMENE) (2020). Cytomegalovirus, Epstein-Barr virus, and human herpesvirus-6 infections in patients with myalgic ?ncephalomyelitis/chronic fatigue syndrome, Journal of Medical Virology, 92, 12, 3682?-3688.

[4] Cook, J., Kleinschmidt, I., Schwabe, C., Nseng, G., Bousema, T., Corran, P. H., Riley, E. M. and Drakeley, C. J. (2011). Serological markers suggest heterogeneity of effectiveness of malaria control interventions on Bioko Island, Equatorial Guinea, Plos One, 6, 9, e25137.

[5] Hsiang, M. S., Hwang, J., Kunene, S., Drakeley, C., Kandula, D., Novotny, J., Parizo, J., Jensen, T., Tong, M., Kemere, J., Dlamini, S., Moonen, B., Angov, E., Dutta, S., Ockenhouse, C., Dorsey, G. and Greenhouse, B. (2012). Surveillance for malaria elimination in Swaziland: a national cross-sectional study using pooled PCR and serology, PloS One, 7, 1, e29550.

[6] Helb, D. A., Tetteh, K. K., Felgner, P. L., Skinner, J., Hubbard, A., Arinaitwe, E., Mayanja-Kizza, H., Ssewanyana, I., Kamya, M. R., Beeson, J. G., Tappero, J., Smith, D. L., Crompton, P. D., Rosenthal, P. J., Dorsey, G., Drakeley, C. J., and Greenhouse, B. (2015). Novel serologic biomarkers provide accurate estimates of recent Plasmodium falciparum exposure for individuals and communities, Proceedings of the National Academy of Sciences of the United States of America, 112, 32, E4438?E4447.

[7] Loebel, M., Eckey, M., Sotzny, F., Hahn, E., Bauer, S., Grabowski, P., Zerweck, J., Holenya, P., Hanitsch, L. G., Wittke, K., Borchmann, P., Rüffer, J. U., Hiepe, F., Ruprecht, K., Behrends, U., Meindl, C., Volk, H. D., Reimer, U., and Scheibenbogen, C. (2017). Serological profiling of the EBV immune response in Chronic Fatigue Syndrome using a peptide microarray, PloS One, 12, 6, e0179124.

[8] Lammie, P. J., Moss, D. M., Brook Goodhew, E., Hamlin, K., Krolewiecki, A., West, S. K. and Priest, J. W. (2012). Development of a new platform for neglected tropical disease surveillance, International Journal for Parasitology, 42, 9, 797?-800.

[9] Blomberg, J., Rizwan, M., Böhlin-Wiener, A., Elfaitouri, A., Julin, P., Zachrisson, O., Rosén, A. and Gottfries, C. G. (2019). Antibodies to Human Herpesviruses in Myalgic Encephalomyelitis/Chronic Fatigue Syndrome Patients, Frontiers in Immunology, 10, 1946.

[10] Sowa, M., Hiemann, R., Schierack, P., Reinhold, D., Conrad, K., and Roggenbuck, D. (2017). Next-Generation Autoantibody Testing by Combination of Screening and Confirmation-the CytoBead ® Technology, Clinical Reviews in Allergy & Immunology, 53,1, 87?-104.

[11] van den Hoogen, L. L., Présumé, J., Romilus, I., Mondélus, G., Elismé, T., Sepúlveda, N., Stresman, G., Druetz, T., Ashton, R. A., Joseph, V., Eisele, T. P., Hamre, K., Chang, M. A., Lemoine, J. F., Tetteh, K., Boncy, J., Existe, A., Drakeley, C. and Rogier, E. (2020). Quality control of multiplex antibody detection in samples from large-scale surveys: the example of malaria in Haiti, Scientific Reports, 10, 1, 1135.

[12] Wu, L., Hall, T., Ssewanyana, I., Oulton, T., Patterson, C., Vasileva, H., Singh, S., Affara, M., Mwesigwa, J., Correa, S., Bah, M., D’Alessandro, U., Sepúlveda, N., Drakeley, C., and Tetteh, K. (2020). Optimisation and standardisation of a multiplex immunoassay of diverse Plasmodium falciparum antigens to assess changes in malaria transmission using sero-epidemiology, Wellcome Open Research, 4, 26.

[13] McLachlan, G. and Peel, D (2000). Finite Mixture Models. John Wiley & Sons, New York.

[14] Gay, N.J. (1996). Analysis of serological surveys using mixture models: application to a survey of parvovirus B19, Statistics in Medicine, 15, 1567–1573.

[15] Chis Ster, I. (2012). Inference for serological surveys investigating past exposures to infections resulting in long-lasting immunity – an approach using finite mixture models with concomitant information, Journal of Applied Statistics, 39, 11, 2523–2542.

[16] Rogier, E., Wiegand, R., Moss, D., Priest, J., Angov, E., Dutta, S., Journel, I., Jean, S. E., Mace, K., Chang, M., Lemoine, J. F., Udhayakumar, V., and Barnwell, J. W. (2015). Multiple comparisons analysis of serological data from an area of low Plasmodium falciparum transmission, Malaria Journal, 14, 436.

[17] Parker, R. A., Erdman, D. D. and Anderson, L. J. (1990). Use of mixture models in determining laboratory criterion for identification of seropositive individuals: application to parvovirus B19 serology, Journal of Virological Methods, 27, 2, 135-?144.

[18] Baughman, A. L., Bisgard, K. M., Lynn, F. and Meade, B. D. (2006). Mixture model analysis for establishing a diagnostic cut-off point for pertussis antibody levels, Statistics in Medicine, 25, 2994–3010.

[19] Rota, M. C., Massari, M., Gabutti, G., Guido, M., De Donno, A. and Ciofi degli Atti, M.L. (2008). Measles serological survey in the Italian population: interpretation of results using mixture model, Vaccine, 26, 34, 4403-?4409.

[20] Nhat, N., Todd, S., de Bruin, E., Thao, T., Vy, N., Quan, T. M., Vinh, D. N., van Beek, J., Anh, P. H., Lam, H. M., Hung, E., Lien, N., Hong, T., Farrar, J., Simmons, C. P., Chau, N., Koopmans, M. and Boni, M. F. (2017). Structure of general-population antibody titer distributions to influenza A virus, Scientific Reports, 7, 1, 6060.

[21] Moreira da Silva, J., Prata, S., Domingues, T. D., Leal, R. O., Nunes, T., Tavares, L., Almeida, V., Sepúlveda, N. and Gil, S. (2020). Detection and modeling of anti-Leptospira IgG prevalence in cats from Lisbon area and its correlation to retroviral infections, lifestyle, clinical and hematologic changes, Veterinary and Animal Science, 10, 100144.

[22] Sepúlveda, N., Stresman, G., White, M.T. and Drakeley, C.J. (2015). Current Mathematical Models for Analyzing Anti-Malarial Antibody Data with an Eye to Malaria Elimination and Eradication, Journal of Immunology Research, 2015, 738030.

[23] Basso, R.M., Lachos, V.H., Cabral, C.R.B. and Gosh, P. (2010). Robust mixture modelling based on scale mixtures of skew-normal distributions, Computational Statistics and Data Analysis, 54, 2926–2941.

[24] Cliff, J.M., King, E.C., Lee, J.S., Sepúlveda, N., Wolf, A.S., Kingdon, C., Bowman, E., Dockrell, H.M., Nacul, L., Lacerda, E. and Riley, E.M. (2019). Cellular Immune Function in Myalgic Encephalomyelitis/Chronic Fatigue Syndrome (ME/CFS), Frontiers in immunology, 10, 796.

[25] Fukuda, K., Straus, S. E., Hickie, I., Sharpe, M. C., Dobbins, J. G., and Komaroff, A. (1994). The chronic fatigue syndrome: a comprehensive approach to its definition and study. International Chronic Fatigue Syndrome Study Group, Annals of Internal Medicine, 121, 12, 953?-959.

[26] Carruthers, B. M., Jain, A. K., De Meirleir, K. L., Peterson, D. L., Klimas, N. G., Lerner, A. M., Bested, A. C., Flor-Henry, P., Joshi, P., Peter Powles, A. C., Sherkey, J. A. and van de Sande, M. I. (2003). Myalgic Encephalomyelitis/Chronic Fatigue Syndrome, Journal of Chronic Fatigue Syndrome, 11, 1, 7–115,

[27] Rasa, S., Nora-Krukle, Z., Henning, N., Eliassen, E., Shikova, E., Harrer, T., Scheibenbogen, C., Murovska, M., Prusty, B. K. and European Network on ME/CFS (EUROMENE) (2018). Chronic viral infections in Myalgic Encephalomyelitis/Chronic Fatigue Syndrome (ME/CFS), Journal of Translational Medicine, 16, 1, 268.

[28] Lacerda, E. M., Bowman, E. W., Cliff, J. M., Kingdon, C. C., King, E. C., Lee, J.S., Clark, T.G., Dockrell, H.M., Riley, E.M., Curran, H. and Nacul, L. (2017). The UK ME/CFS Biobank for biomedical research on Myalgic Encephalomyelitis/Chronic Fatigue Syndrome (ME/CFS) and Multiple Sclerosis, Open Journal of Bioresources, 4, 4.

[29] Lacerda, E. M., Mudie, K., Kingdon, C.C., Butterworth, J.D., O’Boyle, S. and Nacul, L. (2018). The UK ME/CFS Biobank: A Disease-Specific Biobank for Advancing Clinical Research Into Myalgic Encephalomyelitis/Chronic Fatigue Syndrome, Frontiers in Neurology, 9, 1026.

[30] Lin, T.I. and Lee, J.C. and Yen, S.Y. (2007). Finite mixture modelling using the Skew-Normal distribution, Statistica Sinica, 17, 909–927.

[31] Azzalini, A. (1985). A Class of distributions which includes the normal Ones, Scandinavian Journal of Statistics, 12, 171–178.

[32] Azzalini, A. and Capitanio, A. (2003). Distributions generated by perturbation of symmetry with emphasis on a multivariate skew t distribution, J.R.Statist.Soc.B, 65, 367–389.

[33] Lachos Dávila, V. H. and Zeller, C. B. and Cabral, C. R. B. (2018). Finite mixture of skewed distributions, Springer.

[34] Azzalini, A. (2014). The skew-normal and related families, Cambridge University Press.

[35] Ferreira, C.S., Bolfarine H., and Lachos V.H. (2011). Skew scale mixtures of normal distributions: Properties and estimation, Statistical Methodology, 8, 154–171.

[36] Oliveira-Brochado, A. and Martins, F. V. (2005). Assessing the number of components in mixture models: a review, Universidade do Porto, Faculdade de Economia do Porto, 194.

[37] Luko?ien?, O. and Vermunt, J. K. (2009). Determining the number of components in mixture models for hierarchical data, In Advances in data analysis, data handling and business intelligence, 241–249. Springer, Berlin, Heidelberg.

[38] Xie, C. H., Chang, J. Y. and Liu, Y. J. (2013). Estimating the number of components in Gaussian mixture models adaptively for medical image, Optik, 124(23), 6216–6221.

[39] Mehrjou, A., Hosseini, R. and Araabi, B. N. (2016). Improved Bayesian information criterion for mixture model selection, Pattern Recognition Letters, 69, 22–27.

[40] Feng, Z.D and McCullogh, C.E. (1996). Using Bootstrap Likelihood Ratios in Finite Mixture Models, Journal of the Royal Statistical Society, 58, 3, 609–617.

[41] Yu, Y. and Harvill, J.L. (2019). Bootstrap likelihood ratio test for Weibull mixture models fitted to grouped data, Communications in Statistics - Theory and Methods, 48, 18, 4550–4568.

[42] Saraswati, K., Phanichkrivalkosil, M., Day, N. and Blacksell, S. D. (2019). The validity of diagnostic cut-offs for commercial and in-house scrub typhus IgM and IgG ELISAs: A review of the evidence, PLoS Neglected Tropical Diseases, 13, 2, e0007158.

[43] Brent, R.P. (1973). Algorithms for Minimization Without Derivatives, Prentice-Hall, Englewood Cliffs, New Jersey, 73–76.

[44] Prates, M.O., Lachos, V.H. and Cabral, C. (2013). Fitting finite mixture of scale mixture of skew-normal distributions, Journal of Statistical Software, 54, 1–20.

[45] Wolodzko, T. (2020). Additional Univariate and Multivariate Distributions, R CRAN, https://cran.r-project.org/web/packages/extraDistr/index.html.

[46] Azzalini, A. (2020). The Skew-Normal and Related Distributions Such as the Skew-t, R CRAN, https://cran.r-project.org/web/packages/sn/sn.pdf.

[47] Braun, D. K., Dominguez, G. and Pellett, P. E. (1997). Human herpesvirus 6, Clinical Microbiology Reviews, 10, 3, 521-?567.

[48] Su, S. (2007). Fitting Single and Mixture of Generalized Lambda Distributions to Data via Discretized and Maximum Likelihood Methods: GLDEX in R, Journal of Statistical Software, 21, 9, 1–17.

[49] Su, S. (2011). Maximum Log Likelihood Estimation using EM Algorithm and Partition Maximum Log Likelihood Estimation for Mixtures of Generalized Lambda Distributions, Journal of Modern Applied Statistical Methods, 10, 2, 17.

[50] Ramberg, J. and Schmeiser, B. (1974). An Approximate Method for Generating Asymmetric Random Variables, Communications of the Association for Computing Machinery, 17, 78?-82.

[51] Freimer, M., Mudholkar, G., Kollia, G. and Lin, C. (1988). A Study of the Generalised Tukey Lambda Family, Communications in Statistics ? Theory and Methods, 17, 3547?-3567.

[52] McLachlan, G. and Lee, S. (2013). EMMIXuskew: An R Package for Fitting Mixtures of Multivariate Skew t Distributions via the EM Algorithm, Journal of Statistical Software, 55, 12, 1–22.

[53] Ridge, S. E. and Vizard, A. L. (1993). Determination of the optimal cutoff value for a serological assay: an example using the Johne’s Absorbed EIA, Journal of Clinical Microbiology, 31, 5, 1256-?1261.

[54] Kafatos, G., Andrews, N. J., McConway, K. J., Maple, P. A., Brown, K. and Farrington, C. P. (2016). Is it appropriate to use fixed assay cut-offs for estimating seroprevalence?, Epidemiology and infection, 144, 4, 887-?895.

[55] Migchelsen, S. J., Martin, D. L., Southisombath, K., Turyaguma, P., Heggen, A., Rubangakene, P. P., Joof, H., Makalo, P., Cooley, G., Gwyn, S., Solomon, A. W., Holland, M. J., Courtright, P., Willis, R., Alexander, N. D., Mabey, D. C. and Roberts, C. H. (2017). Defining Seropositivity Thresholds for Use in Trachoma Elimination Studies, PLoS Neglected Tropical Diseases, 11, 1, e0005230.

[56] Bouman, J. A., Bonhoeffer, S. and Regoes, R. R. (2020). Estimating seroprevalence with imperfect serological tests: exploiting cutoff-free approaches. bioRxiv, doi: https://doi.org/10.1101/2020.04.29.068999.

